# Pragmatic vs. naïve genetic instrument selection in Mendelian randomization studies: a practical guide

**DOI:** 10.64898/2026.08.19.26359587

**Authors:** Andrew Mason, Giulia Ballabio, Valentina Paz, Reecha Sofat, Victoria Garfield

## Abstract

Mendelian randomization (MR) is widely used to infer causal relationships using genetic variants as instrumental variables, yet the selection of genetic instruments is not always given sufficient attention. Many MR studies rely on default linkage disequilibrium (LD) clumping parameters (*r*^2^ <0.001, 10,000 kb), as implemented in commonly used tools, without assessment of their suitability for specific exposures. We investigated whether this approach yields optimal instruments or whether a more pragmatic strategy yields stronger instruments. Using UK Biobank data, we examined three distinct exposure types—circulating amino acids, body mass index (BMI), and major depressive disorder (MDD). For each phenotype, we systematically varied LD clumping thresholds (*r*^2^ and genomic distance) and evaluated each instrument via both their average strength (*F*-statistic) and total strength (*R*^2^). Across all phenotypes, optimal instruments differed from default parameters and varied by exposure. For amino acids and BMI, more stringent LD thresholds (*r*^2^=0.00001) combined with larger clumping windows improved instrument strength, whereas for MDD, a highly polygenic, binary trait, smaller windows with stringent *r*^2^ maximized variance explained while maintaining *F*-statistics above the desired threshold (>10). Notably, increasing the number of SNPs did not consistently improve instrument quality, highlighting a trade-off between instrument strength and potential pleiotropy. We demonstrate that universal reliance on default LD clumping parameters can lead to suboptimal instruments. We propose a pragmatic framework for instrument selection based on empirical evaluation of strength metrics, improving the robustness and transparency of MR analyses across different exposure types.

**Author summary:** Mendelian randomisation (MR), so-called ‘nature’s randomised control trial’, is a genetic epidemiology tool which exploits the randomisation inherent in the genotypes of individuals to try to establish potential causal links between exposures and outcomes in a variety of contexts. Leveraging population-based genetic data, two-sample Mendelian randomisation combines the associations between genetic variants and exposures (e.g. fasting glucose) in one cohort with the associations between these same variants and an outcome in another (e.g. the risk of a major adverse cardiovascular event).

A crucial step in selecting these genetic variants, is to ensure that they are not in linkage disequilibrium (correlated) with one another (which would violate the core assumption of MR that variants are randomly inherited at conception). Many tools and packages in contemporary programming languages have been developed to perform this crucial step. However, many authors resort to the default options that are pre-determined by these packages. Here, we demonstrate that results and interpretations therein can vary substantially as a function of the choice of parameters while also satisfying the requirements of MR in terms of statistical power and violation of core assumptions. We propose that researchers take a pragmatic approach to genetic instrument selection in MR studies.

## Introduction

Mendelian randomization (MR) has become a popular method to examine causal associations between modifiable exposures and outcomes, by using common genetic variants (single nucleotide polymorphisms -SNPs-) as instrumental variables (1). As we have previously written, genetic instrument selection is an essential part of designing a robust MR study and it should be given due consideration (2,3). Genetic instruments can be selected using a more biological or statistical approach, depending on the exposure under study. It is important to select the best possible instrument to avoid weak instrument bias, which refers to when the instrument is not strongly associated with the exposure (4).

In the last few years, the number of MR studies published has grown exponentially, with more than 3500 published in 2023 and more than 6000 in 2024. It has recently become widely accepted that in MR studies researchers select their genetic instruments based on certain default parameters provided in commonly used MR software packages (e.g., TwoSampleMR R package) (5,6). After an initial screening based on genome-wide significance (p<5*10^-08^), it is standard to perform linkage disequilibrium (LD) clumping to obtain near-independent SNPs. The LD threshold refers to the correlation (r^2^) and thus, variants above the chosen r^2^ will be excluded within the selected distance (kilobases -kb-). In the popular TwoSampleMR R package the default LD clumping parameters are *r*^2^=0.001 within a distance of 10,000kb and these have become widely adopted in applied MR papers.

In cases where the observed data are available for the exposure researchers can and should check that their chosen genetic instrument is strong in terms of average and total strength (2,3). The average strength, determined via the F-statistic should be >10 to avoid weak instrument bias, whereas the R^2^ (amount of variance explained in the exposure by the genetic instrument) helps determine the statistical power of the study. Where the individual-level data are not available, at least F can be approximated using the ‘t-statistic’ method (2,3).

While using the LD clumping parameters described above can lead to selection of the strongest genetic instrument in some scenarios, there is no rationale for why they should be used in all MR studies. We believe that this naïve form of genetic instrument selection MR may have sometimes resulted in suboptimal genetic instruments and that researchers should opt for a more pragmatic approach. In this paper, we aimed to explore whether this current widely adopted strategy for genetic instrument selection in MR studies is robust or whether a more pragmatic approach should be used. We did this by providing empirical examples in the UK Biobank for distinct phenotypes: amino acids, body mass index (BMI) and major depressive disorder (MDD). Our aim was to exemplify our approach using a commonly instrumented continuous exposure (BMI), a more novel set of continuous biomarker exposures (amino acids), and a commonly instrumented binary exposure (MDD). We conclude by providing practical guidance on how to best pragmatically select genetic instruments in MR studies of non-drug exposures.

## Description of the Method

### Empirical examples in the UK Biobank: our hunt for the best genetic instruments

In the examples described below we were interested in finding the best genetic instruments for a biomarker example (amino acids), a metabolic measure (BMI) and a mental health disorder (MDD). We aimed to understand and demonstrate whether in fact genetic instruments for different phenotypes should all be selected using the same approaches (i.e., default parameters provided by MR packages) or whether a pragmatic, empirical approach would work better.

### Sample

The UKB is a cohort of ∼500,000 adults recruited across the UK general population, aged 40-69 years at baseline (2006-2010), details of which are published elsewhere (73). We applied a basic genetic quality control (QC) procedure to the imputed data. At the individual level, we excluded participants with *i)* excessive or minimal heterozygosity, *ii)* more than 10 putative third-degree relatives, *iii)* no consent to extract DNA, *iv)* mismatches between self-reported and genetic sex, *v)* any missing QC information, and *vi*) non-European ancestry (based on a comparison between self-reported and principal component analysis-derived ancestry). We had the following analytical sample sizes: n=378,078 maximum participants for amino acids, n= 461,761 for BMI, n= 371,931 for MDD. These numbers reflect individuals who had both genetic and phenotypic data available. The UKB received ethical approval from the North West Multicentre Research Ethics Committee and obtained informed consent from participants.

#### Phenotypes

##### Branched chain, essential and aromatic amino acids

To date, these biomarkers have not been widely used in MR studies (unlike BMI and MDD). We downloaded the full summary statistics from a recent GWAS by Karjalainen et al. (2024) of random circulating metabolites (7), restricting ourselves to SNP --> X associations for Histidine, Isoleucine, Leucine, Phenylalanine, Tyrosine, and Valine.

##### BMI

For BMI, we downloaded the full summary statistics from a GWAS meta-analysis of body mass and height (8), which contains data from ∼700,000 individuals of European ancestry. This GWAS meta-analysed associations from the Genetic Investigation of ANthropometric Traits Consortium (GIANT) with UK Biobank participants.

##### MDD

We downloaded the full summary statistics from the Howard et al., MDD GWAS (9). Briefly, this meta-GWAS of 807,553 Europeans included data from the Psychiatric Genomics Consortium (PGC (10)), UKB (11) and 23andMe (12). The depression phenotype was operationalised to broadly include MDD clinical diagnoses, self-reports of physician-diagnosed depression, and symptoms of depression measures which were harmonised across cohorts.

#### Pragmatic SNP selection approach

Analyses were performed in PLINK 2.0 and RStudio (version 4.3.1).

We performed LD clumping by constructing a grid of possible parameters for correlation (*r*^2^) and distance (kb) where we tested a total of 5X5 combinations for each phenotype (Figure 1). We used data from the 1000 Genomes Project Northern Europeans from Utah (CEU) cohort as a reference panel. We regressed the respective phenotype (AAs, BMI and MDD) on each genetic instrument to obtain the instrument *R*^2^ and average *F*-statistic to pragmatically guide our choice of which instrument to take forward in future MR analyses.

**Figure 1.**
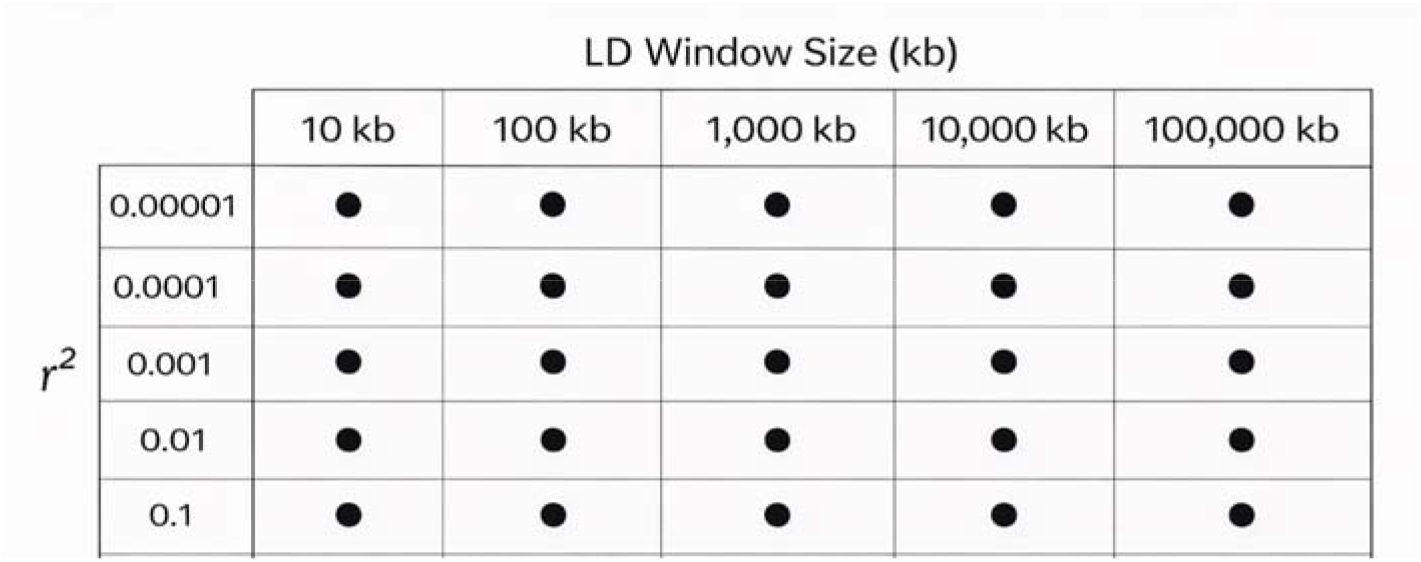
Grid of all combinations tested to generate the strongest instrument for amino acids, BMI and MDD

## Applications

### Strongest genetic instruments for amino acids, BMI and MDD obtained via pragmatic SNP selection

Table 1 presents the strongest instrument for each phenotype, which was selected pragmatically after testing different possible combinations of LD clumping (for all combinations see Supplementary Tables 1-3). Across all three phenotypes we selected the strongest instrument by maximising and balancing both the *F*-statistic and *R*^2^, as they provide information on the average and total strength, respectively. The instruments presented in Table 1 all had *F* >10 (4), indicating that substantial weak instrument bias is unlikely, while R^2^ values varied depending on the phenotype. We also considered LD parameters when choosing the strongest instrument and, in some cases, a less stringent *r*2 and distance (kb) was important for genetic instrument optimisation.

**Table 1.** Details of the strongest genetic instruments for each phenotype.

| Phenotype | $F$ | $R^2$ | Clumping parameters ($r^2$ / kb) | $N_{\text{SNPs}}$ |
| --- | --- | --- | --- | --- |
| *Amino acids* |  |  |  |  |
| Tyrosine | 249.4 | 0.02 | 0.00001 / 100000 | 17 |
| Valine | 150.9 | 0.01 | 0.00001 / 100000 | 12 |
| Phenylalanine | 123 | 0.009 | 0.001 / 100000 | 16 |
| Leucine | 101.4 | 0.008 | 0.00001 / 100000 | 15 |
| Histidine | 74.2 | 0.006 | 0.00001 / 100000 | 15 |
| Isoleucine | 47.5 | 0.006 | 0.00001 / 100000 | 24 |
| *BMI* | 46 | 0.07 | 0.00001 / 1000 | 712 |
| *MDD* | 15 | 0.022 | 0.00001 / 10 | 673 |

*Note.* Instruments are ordered by average ( $F$ -statistic) and total strength ( $R^2$ ).

The strongest amino acid instrument in terms of average and total strength was for Tyrosine, while the weakest instrument produced was for Isoleucine. However, across all instruments the phenotypic variance explained ranged from 0.6% to 2%. Tyrosine (a non-essential amino acid) had the strongest instrument with 17 SNPs, while Isoleucine (an essential amino acid) had the weakest instrument, with 24 SNPs.

For BMI, our optimal instrument explained 7% of the variance with *F* = 46. BMI presented us with much greater choice for the best genetic instrument, as compared with amino acids or MDD. Therefore, we did not select the instrument with the largest F-statistic but instead maximised the R^2^ while also ensuring that the *F* was substantially >10 and that the LD clumping thresholds retained independent variants.

For MDD, the strongest instrument explained 2.2% of the variance with *F* = 15.1. In contrast to BMI and amino acids, MDD provided substantially fewer strong instruments across clumping thresholds. Thus, we prioritised maximising the variance explained while ensuring that the F-statistic remained >10. This meant we selected an instrument with 673 SNPs, which retained sufficient instrument strength while capturing the largest proportion of variance in MDD.

## Discussion

The expanding accessibility of large-scale biobanks and population-based genomic resources, together with the widespread availability of analytical software, has markedly increased the number of MR studies, but it has also exposed important methodological limitations, with critical flaws identified across many published studies (13). In particular, the default LD clumping parameters implemented in the TwoSampleMR R package have been widely adopted without systematic methodological evaluation (13). Here, we assessed whether the prevailing strategy for genetic instrument selection based on LD clumping default parameters in MR is sufficiently robust or whether a more pragmatic and context-dependent approach may be warranted.

Using empirical examples across distinct phenotypes—including amino acids, BMI, and MDD—we compared total (*R*^2^) and average strength (*F*-statistic) metrics under different combinations of LD clumping thresholds (r^2^ and kb). These analyses provide a practical framework for critically appraising instrument selection strategies and highlight the importance of explicitly justifying LD clumping choices in MR studies. Our findings indicate that a more pragmatic, empirically driven approach to instrument selection outperforms the routine use of default LD clumping parameters (*r*^2^=0.001 and 10,000 kb). Importantly, the impact of clumping thresholds differed across phenotypes and according to exposure type.

For all amino acids —except phenylalanine—the combination of *r*^2^ = 0.00001 and a 100,000 kb window yielded the strongest instruments. In contrast, for phenylalanine, the optimal configuration was *r*^2^ = 0.001 with a 100,000 kb window. When considering both the F-statistic and R^2^, tyrosine exhibited the strongest instrument despite comprising 17 SNPs, whereas isoleucine showed the weakest instrument with 24 SNPs. These results demonstrate that a larger number of genetic variants is not always better (2,3). It is therefore crucial not to assume that the largest available GWAS will automatically provide the optimal instrument for a given exposure. Instead, careful consideration should be given to the robustness of the identified SNPs. Selecting a large number of variants could lead to a larger total strength (R^2^) but a weaker average strength (*F*-statistic) and a greater chance of including pleiotropic variants. On the contrary, fewer variants will lead to a lower *R*^2^ but potentially a higher F-statistic, which could result in an instrument with insufficient power (14). A careful balance is therefore required: enough genetic variants should be retained to ensure adequate statistical power and enable the application of more robust methods, such as MR-Egger regression, while avoiding including so many variants that horizontal pleiotropy becomes unavoidable. These results emphasise that instrument quality, rather than sheer quantity, should guide parameter selection.

As with amino acids, BMI—which is also a continuous trait, albeit more widely studied—showed that the combination of *r*^2^ = 0.00001 and a 100,000 kb window yielded the strongest instrument. Again, a larger number of SNPs did not guarantee a better instrument: the tyrosine instrument, comprising 17 SNPs, outperformed the BMI instrument with 712 SNPs in terms of the *F*-statistic, but not *R*^*2*^. For MDD, a binary exposure, the optimal combination was *r*^2^ = 0.00001 and a 10kb window. MDD is known to be highly polygenic, with thousands of variants that contribute very small effects, which is also the case for other psychiatric traits (10). To maximise variance explained (total strength) but maintain adequate average instrument strength, we prioritised inclusion of a larger number of genetic variants using a stringent LD threshold (*r*^2^), rather than a large clumping window as this would substantially reduce instrument strength and subsequent statistical power in downstream MR analyses. It is also important to acknowledge the distinction between *cis* and *trans* acting genetic variants. The former refers to variants within or close to a target gene, while the latter are variants outside of (and often far away from) a target gene. The genetic variants used in the examples presented in this paper are, therefore, *trans* variants.

In MR studies, it is well established that continuous measures should be used where possible, because there are important methodological challenges in the genetic instrumentation of binary exposures (2,15). When instrumenting binary exposures, it is essential to recognise that the analysis implicitly models an underlying continuous liability. Accordingly, MR estimates derived from binary exposures should be interpreted in terms of genetic liability rather than the observed disease status per se (16). Evidence to support this comes from a previous methodological guide we published for careful instrumentation of glycaemic phenotypes. A continuous glycated haemoglobin (HbA1c) instrument explained 3% of the variance with an *F* = 164, while the diabetes instrument explained 1.5% and had an *F* = 27 (2).

## Conclusions

In summary, our findings indicate that the widespread use of the default LD clumping parameters implemented in the TwoSampleMR R package should not be adopted uncritically. Across diverse exposure types, we demonstrate the importance of evaluating clumping parameters according to their impact on instrument strength metrics. We therefore recommend a more reflective and empirically informed approach to genetic instrument selection, recognising that this is one of the most important first steps in MR study design. Universal reliance on default settings is unlikely to be appropriate; instead, clumping decisions should be explicitly justified and instrument strength metrics transparently reported. Overall, this work provides practical guidance for selecting high-quality genetic instruments across exposure types, thereby supporting more rigorous and transparent MR research and strengthening causal inference. Nevertheless, as MR is not a panacea, replication, triangulation of evidence, and complementary approaches—for example, target trial emulation and counterfactual-based methods—remain essential to enhance the robustness and interpretability of findings.

## Data Availability

All data used in the present study are available to bona fide researchers via an application to the UK Biobank

https://www.ukbiobank.ac.uk/

## Acknowledgements

This work is funded by Diabetes Research & Wellness Foundation, Professor David Matthews Non-Clinical Fellowship to VG (ref: SCA/01/NCF/22). RS is an NIHR Research Professor, NIHR303160 is funded by the NIHR for this research project. The views expressed in this publication are those of the author(s) and not necessarily those of the NIHR, NHS or the UK Department of Health and Social Care.

## Supporting information

**Table S1. Amino acid SNPs**

**Table S2. BMI SNPs**

**Table S3. MDD SNPs**

